# Genomics-based identification of a potential causal role for acylcarnitine metabolism in depression

**DOI:** 10.1101/2021.10.18.21265157

**Authors:** Yuri Milaneschi, Matthias Arnold, Gabi Kastenmüller, Siamak Mahmoudian Dehkordi, Ranga R. Krishnan, Boadie W. Dunlop, A. John Rush, Brenda W. J. H. Penninx, Rima Kaddurah-Daouk, for the Mood Disorders Precision Medicine Consortium (MDPMC)

## Abstract

**Background:** Altered metabolism of acylcarnitines – transporting fatty acids to mitochondria – may link cellular energy dysfunction to depression. We examined the potential causal role of acylcarnitine metabolism in depression by leveraging genomics and Mendelian randomization.

**Methods:** Summary statistics were obtained from large GWAS: the Fenland Study (N= 9,363), and the Psychiatric Genomics Consortium (246,363 depression cases and 561,190 controls). Two-sample Mendelian randomization analyses tested the potential causal link of 15 endogenous acylcarnitines with depression.

**Results:** In univariable analyses, genetically-predicted lower levels of short-chain acylcarnitines C2 (Odds Ratio [OR] 0.97, 95% Confidence Intervals [CIs] 0.95-1.00) and C3 (OR 0.97, 95%CIs 0.96-0.99) and higher levels of medium-chain acylcarnitines C8 (OR 1.04, 95%CIs 1.01-1.06) and C10 (OR 1.04, 95%CIs 1.02-1.06) were associated with increased depression risk. No reverse potential causal role of depression genetic liability on acylcarnitines levels was found. Multivariable analyses showed that the association with depression was driven by the medium-chain acylcarnitines C8 (OR 1.04, 95%CIs 1.02-1.06) and C10 (OR 1.04, 95%CIs 1.02-1.06), suggesting a potential causal role in the risk of depression. Causal estimates for C8 (OR=1.05, 95%CIs=1.02-1.07) and C10 (OR=1.05, 95%CIs=1.02-1.08) were confirmed in follow-up analyses using genetic instruments derived from a GWAS meta-analysis including up to 16,841 samples.

**Discussion:** Accumulation of medium-chain acylcarnitines is a signature of inborn errors of fatty acid metabolism and age-related metabolic conditions. Our findings point to a link between altered mitochondrial energy production and depression pathogenesis. Acylcarnitine metabolism represents a promising access point for the development of novel therapeutic approaches for depression.

## INTRODUCTION

Depression is a major public health burden, due to its high prevalence, early onset, chronic nature, and unsatisfactory levels of treatment response (1). Developing more effective treatments requires a better comprehension of depression pathophysiology, which so far remains substantially elusive.

Defects in cellular metabolism and mitochondrial function have been proposed as a potential pathophysiological mechanism in depression.(2–5) Cell metabolism dysfunctions are involved in brain processes (neurotoxicity, impaired neuroplasticity) related to neuropsychiatric diseases including depression and schizophrenia,(6,7) and linked to inflammation and metabolic conditions (e.g. insulin resistance) commonly found in depression.(2,8)

Acylcarnitines (ACs) are biogenic compounds formed from the transfer of the acyl group of a fatty acyl-Coenzyme A to carnitine, and serve as central cogs in the mitochondrial machinery. Dysregulation of AC metabolism has been linked to impaired β-oxidation of fatty acids, thereby reducing mitochondrial energy production.(9,10) Recently, depression has been linked to AC metabolism through studies (11,12) showing altered circulating AC levels in subjects with Major Depressive Disorder as compared to healthy controls. A recent metabolomics study(13) on >1,000 subjects identified lower levels of the medium-chain acylcarnitines decanoylcarnitine (C10) and dodecanoylcarnitine/laurylcarnitine (C12) in participants with elevated depressive symptoms as compared to controls. Furthermore, ACs profiles have been shown to correlate with depression severity and variation in response to treatment with SSRIs and Ketamine (14–16).

Despite this preliminary evidence, whether the observed alterations of AC levels have a role in depression onset or are a consequence of the disease is unknown. The few available and mainly cross-sectional observational studies preclude making any causal inferences because observational associations may emerge as a result of (1) residual confounding from several biopsychosocial factors such as physical inactivity, poor dietary habits, or medication use, impacting AC levels and depression, or (2) reverse causation, with subclinical depression influencing AC levels via behavioral and lifestyle-related (e.g. reduction in physical activity, worsening of dietary habit) mechanisms secondary to the ensuing disorder.

Genomics provides unique opportunities to investigate causality between traits applying new statistical tools such as Mendelian randomization (MR)(17) to results from large genome-wide association studies (GWAS). MR is a technique inferring causality by leveraging genetic variants as proxy instruments, which are less likely related to confounders due to random allele segregation and not reversely affected by the phenotype. Previous metabolite GWAS (mGWAS) (18) showed that a limited number of genetic variants explained a large proportion circulating levels of ACs, a pattern reflecting a sparse genetic architecture with few biologically relevant loci harboring key genes for AC synthesis, metabolism and transport. This set of genetic variants with clear biological connotation and large effect sizes provide valuable instruments to examine the potential causal role of AC metabolism in the development of depression. In the present study, we therefore used results from recent large GWAS and applied MR analyses exploring the potential bidirectional causality between AC levels and depression.

## METHODS

### GWAS summary statistics and instruments selection

Summary statistics were obtained from large GWAS of international consortia. Summary statistics for ACs measured with a targeted metabolomics platform (Biocrates AbsoluteIDQ p180 Kit; Biocrates Life Sciences AG, Innsbruck, Austria) were retrieved from a mGWAS in 9,363 samples from the Fenland study,(19) a population-based cohort of adults recruited from general practice lists in Cambridgeshire, UK (interrogation of the mGWAS results can be performed at www.omicscience.org). The Psychiatric Genomics Consortium performed an overarching meta-analysis (20) of all available GWAS datasets with depression phenotypes (hereafter labeled as DEP) including established Major Depressive Disorder diagnosis or self-declared depression, totaling 246,363 cases and 561,190 controls.

AC summary statistics (∼10M SNPs, 1000Genomes phase 3 imputation panel) were processed by removing strand ambiguous SNPs and with MAF<1%. Variants overlapping with those reported in depression GWAS (∼8M SNPs, 1000Genomes phase 3 imputation panel) were clumped (10,000 kb window, *r*^*2*^=0.01, EUR population of 1000Genomes used as linkage disequilibrium reference) to identify independent significantly (p<5.0e-8) associated SNPs. For the present analyses, we retained data from 15 ACs with at least two independent associated SNPs (eTable 1).

### Genetic architecture of selected acylcarnitines

Full GWAS summary statistics of ACs were used to derive two parameters related to their genetic architecture. SNP-heritability (*h*^*2*^_*SNP*_, the proportion of trait variance explained by the joint effect of all genotyped common SNPs) was estimated using linkage-disequilibrium score regression (LDSC).(21) Pairwise genetic correlations (*rg*, determined by the number of SNPs and their level of concordance shared between two traits) between ACs were estimated using the high-definition likelihood (HDL) method recently developed(22) as an extension of bivariate LDSC.(21)

### Mendelian randomization analyses

Two-sample Mendelian randomization (2SMR) analyses (23) based on GWAS summary statistics were performed to test the potential causal role of ACs on depression risk. For each AC used as exposure, genome-wide significant independent SNPs were used as instruments. Selected SNPs were aligned on the positive strand for exposures and outcome (DEP). F-statistics (all F>10, eTable 1) indicated that the strength of selected genetic instruments was adequate.(24) Lack of sample overlap across the two discovery GWAS reduced the likelihood of causal estimates biased toward the observational correlation.(25) First, a series of univariable 2SMR analyses were performed based on the inverse variance weighted (IVW) estimator,(26) pooling SNP-exposure/SNP-outcome estimates inversely weighted by their standard error. Since IVW assumes that all SNPs are valid instruments or that the sum of the directional bias is zero, the robustness of significant results was tested in sensitivity analyses based on weighted median and MR-Egger estimators. The weighted median(27) estimator is the median of the weighted empirical distribution function of individual SNP ratio estimates, providing consistent effect estimates even if half of the instruments are invalid. The MR-Egger regression (28) consists of a weighted linear regression similar to IVW relying on the InSIDE assumption (the magnitude of any pleiotropic effects should not correlate with the magnitude of the main effect), providing valid effect estimate even if all SNPs are invalid instruments under the ‘NOME’ assumption (uncertainty in the SNP-exposure association estimates is negligible).(29) At least 10 genetic instruments are recommended(28) to run adequately powered MR-Egger analyses.

Furthermore, heterogeneity among included SNPs was tested via Cochran’s Q test, single SNP, and leave-one-out SNP analyses. The presence of potential horizontal pleiotropy (a genetic instrument for exposure influencing the outcome by mechanisms other than exposure) was tested using the MR-Egger intercept(29) and the MR-PRESSO (pleiotropy residual sum and outlier) method(30) (supplemental methods). Finally, we performed reversed univariable 2SMR analyses testing the potential causal impact of depression liability on ACs circulating levels.

ACs significantly associated at nominal level with DEP risk in univariable analyses were carried forward in multivariable MR (MVMR) analyses,(31) in which the association of each exposure instrument was conditioned on the others.

Finally, we performed follow-up analyses evaluating the impact of the strength of genetic instruments on previous results by repeating all 2SMR analyses for a subset of ACs with stronger instruments derived from larger discovery GWAS summary statistics. For this we meta-analyzed results for the Fenland Study with those from a previous mGWAS meta-analyses (32) of 7,478 samples from seven European cohorts. The overlap among the two datasets were limited to 10 ACs and ∼2M SNPs, due to the use of a different metabolic platform and a less dense imputation panel in the previous mGWAS (supplemental methods for detailed descriptions of the datasets and processing steps).

Analyses were conducted in R v4.0.0 (R Project for Statistical Computing) using the MR-Base package.(33) Statistical significance level was set at α=0.05, two-sided. False discovery rate (FDR) q-values according to the Benjamini-Hochberg procedure were additionally reported in univariable analyses testing separately all 15 ACs.

## RESULTS

Selected data included GWAS summary statistics of 15 metabolites, including free carnitine (C0), and short-chain (C0, C2, C3, C4, C5), medium-chain (C6, C8, C9, C10, C10:1, C12) and long-chain (C14:1, C16, C18:1, C18:2) ACs with at least two independent associated SNPs (median N of associated SNPs = 5; eTable 1).

Figure 1 shows genetic architecture parameters of the selected ACs. The diagonal reports *h*^*2*^_*SNP*,_ which could be estimated as significantly different from 0 for eleven of the fifteen ACs examined. Estimates of *h*^*2*^_*SNP*_ were substantially similar (full results eTable 2) and varied from 0.14 for C2 (se=0.07) and C18:2 (se=0.05) to 0.30 (se=0.05) for C6. The heatmap displays pairwise *rg* between ACs (full results eTable 3). Genetic correlations were moderate (across chain lengths) to strong (within similar chain lengths subgroups), in particular within the medium-chain ACs, with C8-C10 (*rg*=0.98, se=0.01) and C10-C10:1 (*rg*=0.98, se=0.17) correlations statistically not different from unity.

**Figure 1.**
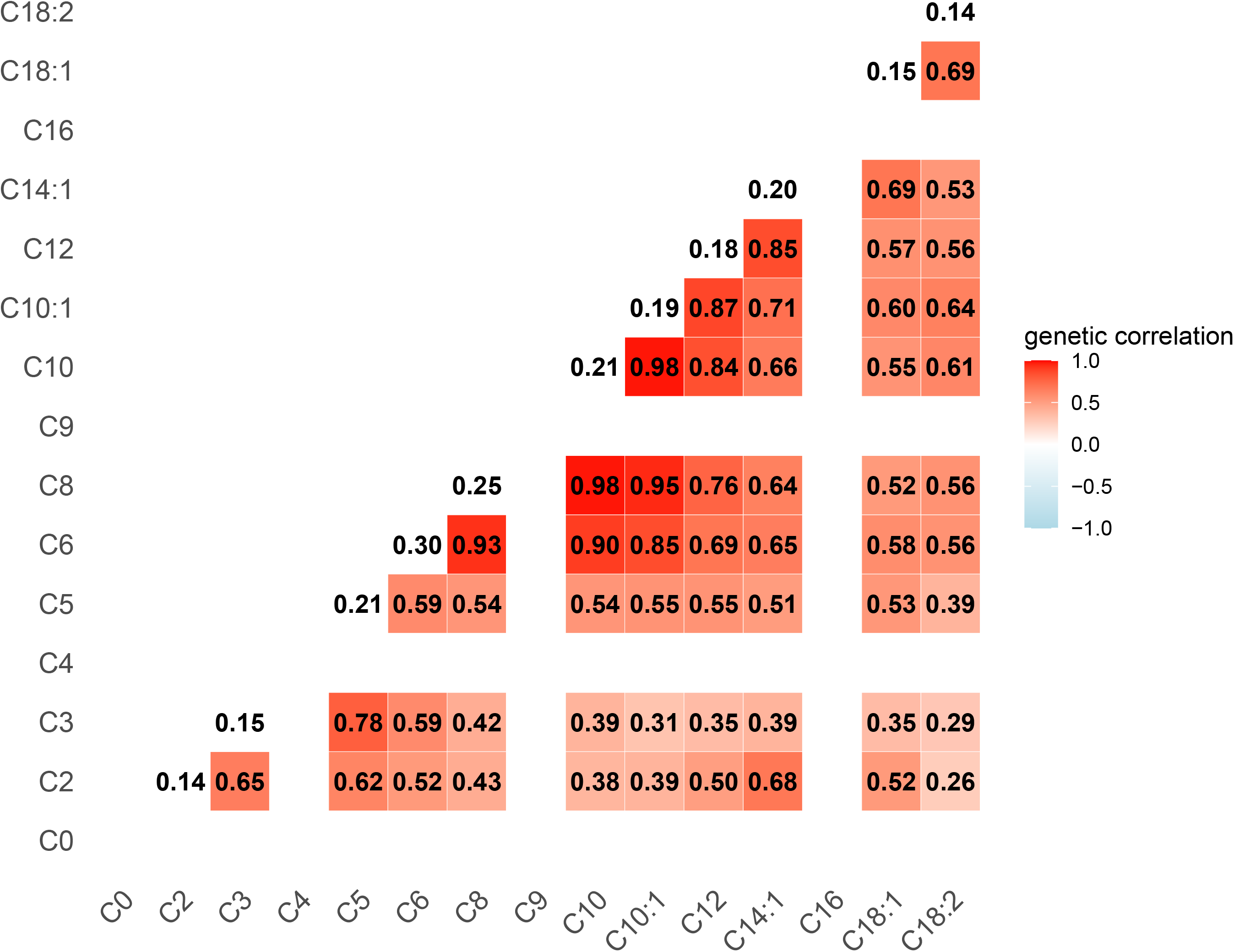
SNP-heritability and pairwise genetic correlation of acylcarnitines. Diagonal: SNP-heritability (*h*^*2*^_*SNP*_) estimates (full results eTable 2). Heatmap: genetic correlation (*rg*) coefficients (full results eTable3).

### Univariable Mendelian randomization analyses

Figure 2 shows the results of univariable 2SMR IVW analyses estimating the risk of DEP (expressed as odds ratios [ORs] and 95% confidence intervals [95%CIs]) per SD increase in genetically-predicted levels of _*(log)*_ACs (full results eTable 4). A lower risk of DEP was significantly associated with genetically-predicted higher levels of the short-chain ACs C2 (OR=0.97, 95%CIs=0.95-1.00, *p*=1.7e-2, *q*=0.06) and C3 (OR=0.97, 95%CIs=0.96-0.99, *p*=1.7e-2, *q*=0.03). Conversely, a higher DEP risk was associated with genetically-predicted higher levels of the medium-chain ACs C8 (OR=1.04, 95%CIs=1.01-1.06, *p*=3.3e-3, *q*=0.02) and C10 (OR=1.04, 95%CIs=1.02-1.06, *p*=6.7e-4, *q*=0.01).

**Figure 2.**
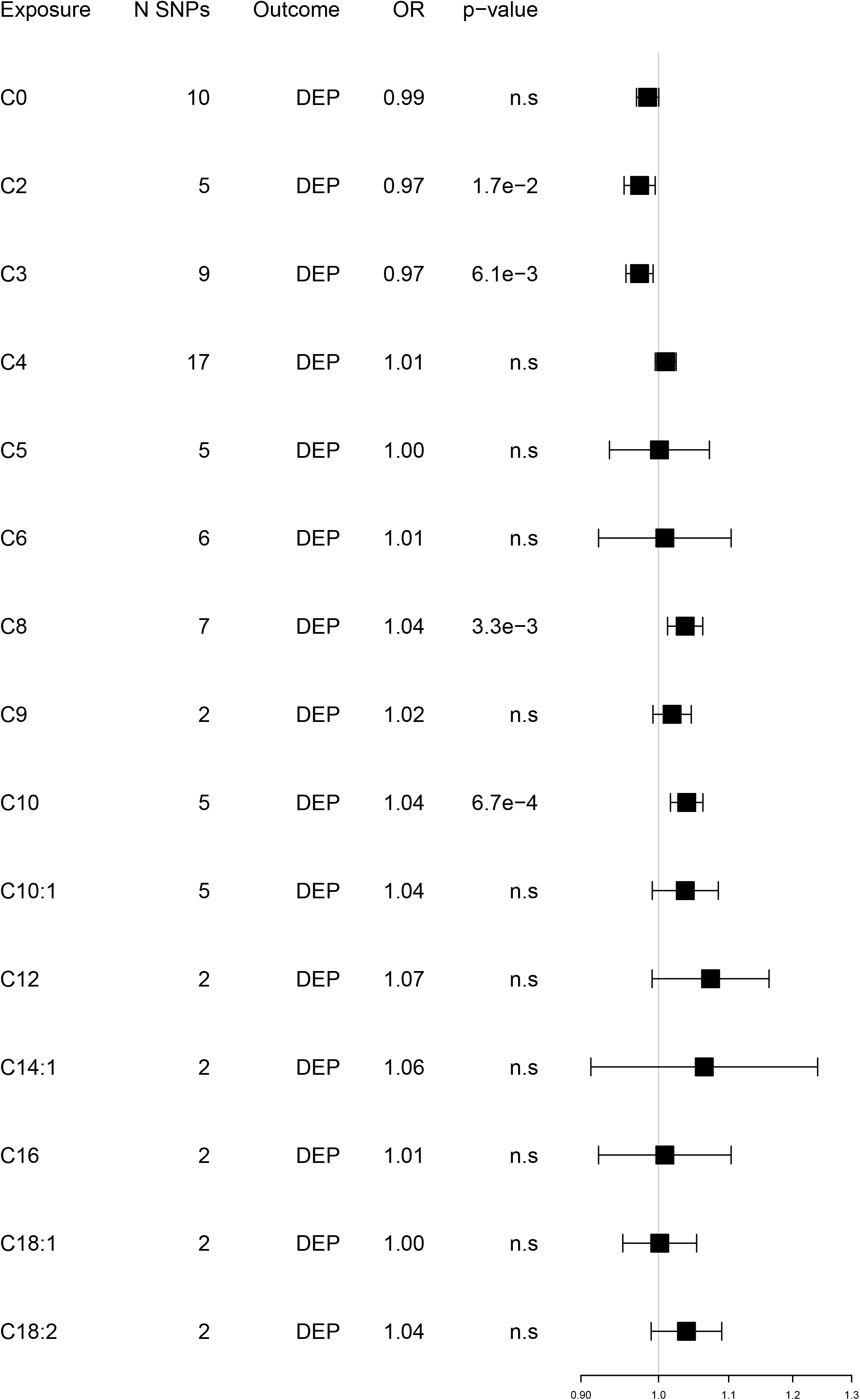
Univariable Mendelian randomization analyses. Exposure: acylcarnitines. Outcome: depression. Odds ratios [ORs] and 95% confidence intervals per SD increase in genetically-predicted levels of _*(log)*_ACs. Full results eTable 4.

Sensitivity analyses confirmed the robustness of these results: causal estimates obtained via weighted-median and MR-Egger estimators were completely in line with those from IVW (Table 1). MR-Egger estimates were not statistically significant, likely due to the limited number of genetic instruments, which were lower than those recommended (N ≥10) to achieve adequate statistical power.(28) Nevertheless, MR-Egger causal estimates were highly convergent with those obtained by other 2SMR analyses. Furthermore, heterogeneity across SNPs was not statistically significant (eTable 5). Inspections of single-SNP and leave-one-out-SNP analyses plots for C8 (eFigure 1-2), which had a Cochran’s Q *p*-value=0.05, did not reveal the presence of outlier SNPs. MR-Egger intercept estimates and results from MR-PRESSO (eTable 6) did not show statistically significant evidence of horizontal pleiotropy. Finally, we performed reversed 2SMR analyses examining the potential causal role of DEP on the levels of the 15 ACs. Figure 3 depicts estimates representing change in SD of _*(log)*_AC levels per doubling (2-fold increase) in the prevalence of the exposure (full results eTable 7). None of the causal estimates were statistically significant, indicating lack of evidence for a reversed causal effect of depression liability on AC levels.

**Table 1.** Association between genetically-predicted levels of four acylcarnitines and risk of depression obtained with different Mendelian Randomization estimators.

| Exposure | N SNPs | Outcome | Inverse Variance Weighted |  |  |  | Weighted median |  |  |  | MR-Egger |  |  |  |
| --- | --- | --- | --- | --- | --- | --- | --- | --- | --- | --- | --- | --- | --- | --- |
|  |  |  | OR | ICI | uCI | *p-value* | OR | ICI | uCI | *p-value* | OR | ICI | uCI | *p-value* |
| C2 | 5 | DEP | 0.97 | 0.95 | 1.00 | 1.7E-02 | 0.97 | 0.95 | 0.99 | 9.0E-03 | 0.97 | 0.93 | 1.02 | 0.39 |
| C3 | 9 | DEP | 0.97 | 0.96 | 0.99 | 6.1E-03 | 0.97 | 0.95 | 0.99 | 8.6E-03 | 0.99 | 0.94 | 1.05 | 0.85 |
| C8 | 7 | DEP | 1.04 | 1.01 | 1.06 | 3.0E-03 | 1.02 | 1.00 | 1.05 | 4.5E-02 | 1.02 | 0.97 | 1.08 | 0.39 |
| C10 | 5 | DEP | 1.04 | 1.02 | 1.06 | 6.7E-04 | 1.03 | 1.01 | 1.06 | 1.3E-02 | 1.04 | 0.98 | 1.10 | 0.28 |

Odds ratios (ORs) and 95% confidence intervals (ICI, lower bound; uCI, upper bound) per SD increase in genetically-predicted levels of $_{(log)}$ ACs

**Figure 3.**
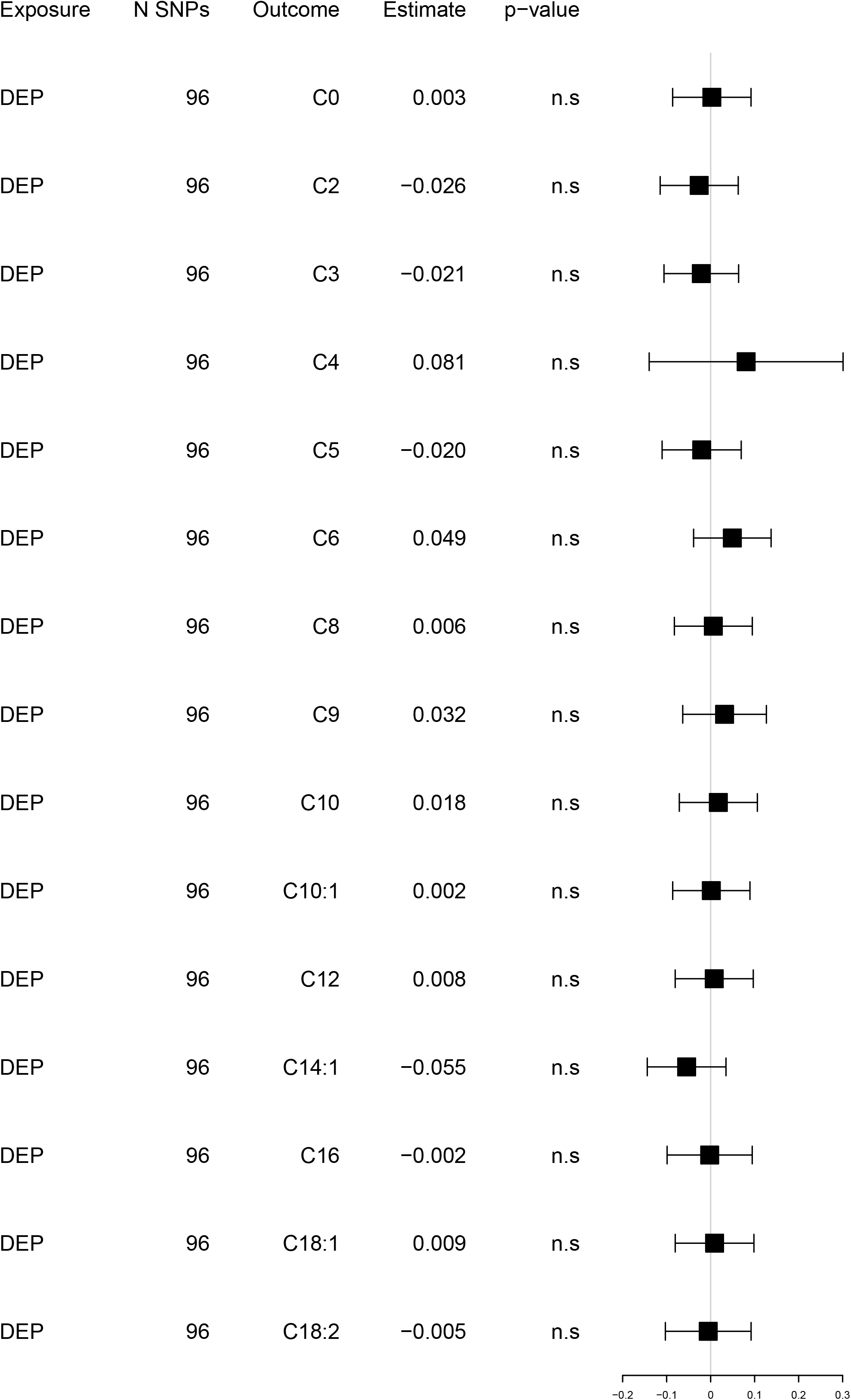
Reversed univariable Mendelian randomization analyses. Exposure: depression. Outcome: acylcarnitines Estimates and 95% confidence intervals of change in SD of _*(log)*_AC levels per doubling (2-fold increase) in the prevalence of the exposure. Full results eTable 7.

### Multivariable Mendelian randomization analyses

Genetic instruments for C2, C3, C8 and C10 were applied in MVMR analyses conditioning the effect of each exposure instrument on the others. Since C8-C10 genetic correlation was equal to 1, we alternatively included them in two models with C2 and C3 (Figure 4, upper and middle panel; full results eTable 8). Both models showed that only genetically-predicted higher levels of the medium-chain ACs, C8 (OR=1.04, 95%CIs=1.02-1.06, *p*=3.5e-5) or C10 (OR=1.04, 95%CIs=1.02-1.06, *p*=5.6e-7), were similarly associated with higher risk of DEP. In a final model including all genetic instruments (Figure 4, lower panel) only that of C10 remained significantly associated (OR=1.16, 95%CIs=1.03-1.31, *p*=0.01) with DEP risk.

**Figure 4.**
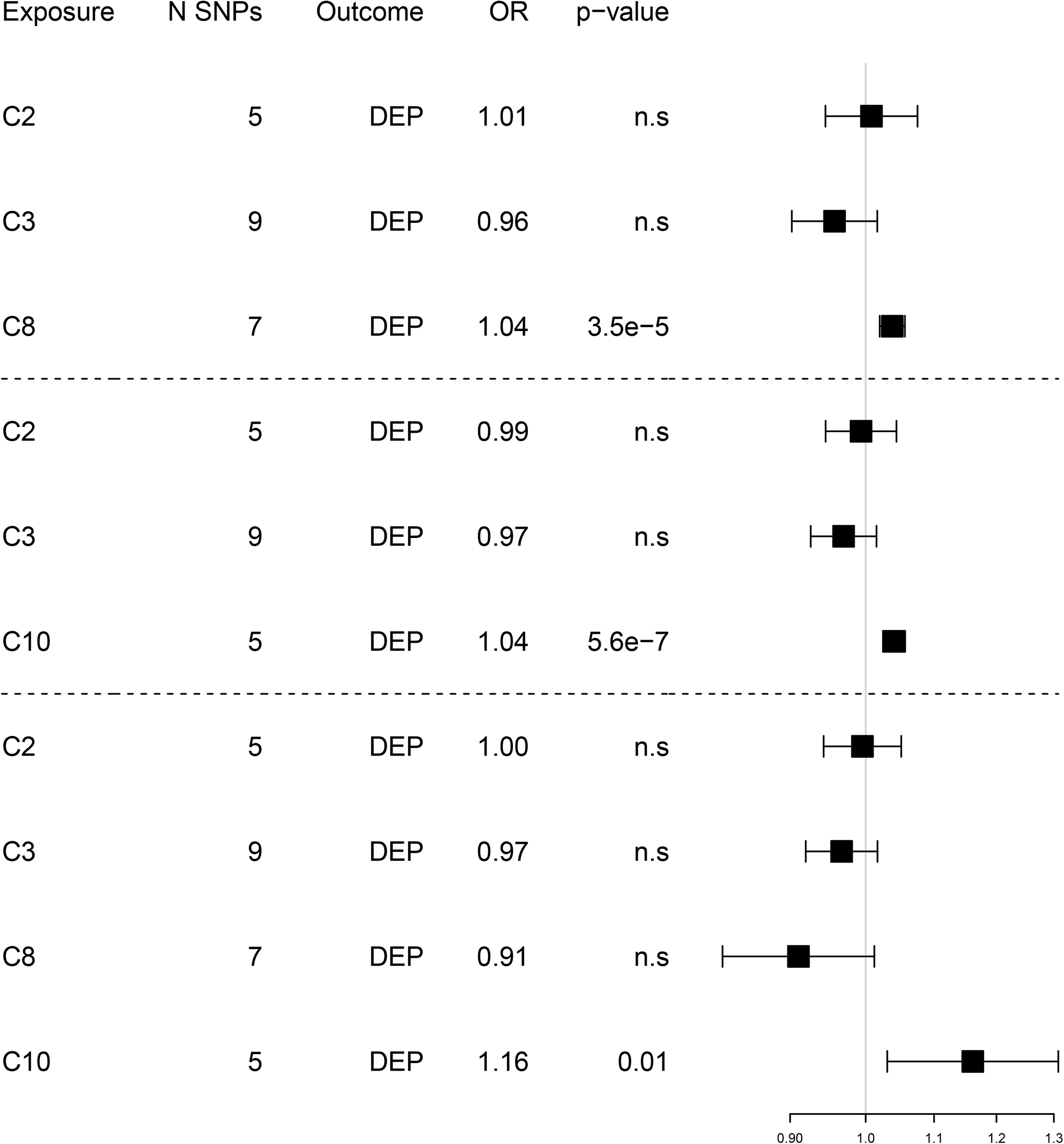
Multivariable Mendelian randomization analyses. Joint exposure: acylcarnitines with statistically significant causal estimates in univariable analyses. Outcome: depression. Odds ratios [ORs] and 95% confidence intervals per SD increase in genetically-predicted levels of _*(log)*_ACs. Full results eTable 8.

### Follow-up analyses

We repeated all 2SMR analyses for a subset of 10 of the 15 metabolites (free carnitine C0; short-chain: C2, C3; medium-chain: C8, C9, C10; C10:1; long-chain: C14:1, C18:1, C18:2) using genetic instruments derived from larger GWAS summary statics, obtained pooling results from the Fenland Study and a previous mGWAS(32) (supplemental methods). The almost doubling in sample size enabled the inclusion of relatively stronger individual instruments but not of a larger number of independently associated SNPs (eTable 9), further supporting the notion of a genetic architecture for ACs characterized by few biologically relevant loci. Univariable 2SMR analyses (eTable 10) confirmed statistically significant causal estimates for C2 (OR=0.96, 95%CIs=0.94-0.99, *p*=1.4e-2), C3 (OR=0.96, 95%CIs=0.93-0.99, *p*=5.6e-3), C8 (OR=1.05, 95%CIs=1.02-1.08, *p*=2.7e-3) and C10 (OR=1.05, 95%CIs=1.00-1.09, *p*=4.0e-2), and revealed a nominal significant association between genetically-predicted levels of C0 and lower risk of DEP (OR=0.98, 95%CIs=0.96-1.00, *p*=1.7e-2). All effect sizes were highly consistent with those obtained in the main analyses, suggesting that the Fenland Study GWAS already provided adequately strong instruments for MR. MVMR analyses (eTable 11) including the abovementioned ACs confirmed that the association with DEP was mainly driven by genetic instruments indexing medium-chain ACs (C8: OR=1.05, 95%CIs=1.02-1.07, *p*=1.4e-4; C10: OR=1.05, 95%CIs=1.02-1.08, *p*=2.1e-3).

## DISCUSSION

This study examined the potential causal role of AC metabolism in depression using recent genetic data and Mendelian randomization analyses. We found that genetically-predicted higher levels of the short-chain acetylcarnitine (C2) and propionylcarnitine (C3) were inversely associated with the risk of depression, while genetically-predicted higher levels of the medium-chain octanoylcarnitine (C8) and decanoylcarnitine (C10) were associated with an increased depression risk. In multivariable analyses, the association with higher depression risk was mainly driven by genetic instruments indexing higher levels of medium-chain ACs. Furthermore, no evidence was found for a potential reversed association between depression liability as exposure and AC levels as outcome.

Present findings showed that circulating ACs are substantially influenced by genetics, with common SNPs explaining 14-30% of the population variance in AC levels (SNP-heritability). This is consistent with previous studies(18) showing large heritability estimates for these metabolites. Furthermore, the different ACs shared a substantial proportion their genetic liability, as indicated by moderate (across chain lengths) to strong (within short, medium, and long chain lengths) genetic correlations between ACs. These patterns of genetic overlap are in line with genetic association results replicated across many mGWASs(18,19,32,34,35). These studies reported chain length-unspecific associations at loci harboring ACs transporters, such as the organic cation transporters *SLC22A4* and *SLC22A5*, or co-enzymes involved in beta-oxidation, such as *ETFDH*. In contrast, enzymes with narrower substrate specificity, such as the short-, medium-, and long-chain acyl-CoA dehydrogenases *ACADS, ACADM*, and *ACADL* that are involved in beta-oxidation, show more specific association patterns for the respective AC species, resulting in the stronger genetic correlations observed here.

The main findings of the present study suggest that higher levels of medium-chain ACs, or the mechanism translating genetic variation to higher levels of these ACs, are potentially causally involved in the development of depression. In a broader perspective, the present findings are consistent with the hypothesis (2) implicating mitochondrial energetic dysfunctions in the pathophysiology of depression. Previous studies(36–38) have shown evidence of impaired mitochondrial respiration and energy output in peripheral tissues of patients with depression. ACs exert a key role in the mitochondria, transporting fatty acids for beta-oxidation and energy production. Plasma or serum AC levels can be used as biomarkers tagging abnormalities in beta-oxidation and inborn errors of metabolism such as MCAD (Medium-chain acyl-coenzyme A dehydrogenase) deficiency, whose clinical screening includes the assessment of potentially elevated levels of octanoylcarnitine (C8) and decanoylcarnitine (C10).(39) Intriguingly, clinical manifestations of fatty acid oxidation disorders include neuropsychological and behavioral symptoms present in depression and other psychiatric disorders, such as cognitive impairments, mood alterations, irritability, sleep and appetite dysregulations and low energy.(40) Recent metabolomics analyses(13) in a population-based sample showed that subjects with elevated depressive symptoms, as compared to healthy controls, had lower levels of medium-chain ACs such as decanoylcarnitine (C10) and dodecanoylcarnitine (C12), a finding that seem in contrast with the present MR results. Nevertheless, discrepancies between observational and genetic estimates are not uncommon (e.g. C-reactive protein levels and schizophrenia (41)) and may be explained by methodological differences across analyses, including for instance the populations selected. Nevertheless, such discrepancies may also hint to relevant dynamics: for instance, it could be speculated that genetically-predicted high levels of medium-chain ACs (MR estimates) index the vulnerability for impaired mitochondrial fatty acid β-oxidation; in contrast, the observed low levels of medium-chain ACs (observational estimates) in depression may be the results of compensatory mechanisms aiming at improving β-oxidation. The potential impact of compensatory mechanisms is in line with recent data(15) showing that antidepressant treatment is associated with increase in short-chain ACs and decrease in medium-chain ACs, suggesting enhanced mitochondrial function. At the current stage such hypothesis remain merely speculative and in order to reconcile genetics and observational estimates further research is needed including longitudinal studies and experimental medicine approaches.

Cellular energetic dysfunctions may be linked to depression through several pathways. In the brain, altered cellular metabolism may lead to neurotoxicity and impaired neuroplasticity.(6,7) In animal models, supplementation with acetylcarnitine (C2), for which higher genetically-predicted concentrations were associated with a reduced risk of depression in the present univariable MR analyses, has been shown to promote neuroplasticity and neurotrophic factor synthesis, to modulate glutamatergic dysfunction and related neuronal atrophy in the hippocampus and amygdala, and to ameliorate depression-like behavioral phenotypes.(42–46) Furthermore, mitochondrial dysfunctions may determine immune system alterations linked to depression. Oxidative stress may trigger the activation of the immune system innate branch by stimulating the release of pro-inflammatory cytokines that, in turn, further stimulate oxidative stress (“autotoxic loop”) and participate in depression pathophysiological processes such as alterations in monoaminergic neurotransmission, tryptophan degradation towards neurotoxic end-products, glutamate-related increased excitotoxicity, decreased neurotrophic factors synthesis or hypothalamic-pituitary-adrenal-axis activity disruption(2,47,48). Furthermore, energy dysfunctions in immune cells may explain the impairments in acquired immunity often observed in depression. Interestingly, recent data (49) on T cells of depressed patients showed an impaired metabolic profile accompanied by increased expression of CTP1a (carnitine palmitoyltransferase 1A), the mitochondrial enzyme responsible for the formation of ACs. Studies based on animal models showed that increased expression of CTP1a are determined by exposure to chronic stress (50) and may trigger(51) hyperphagia with consequent weight gain, symptoms of so-called “atypical” depression(8), hyperglycemia and insulin resistance. Furthermore, cellular energy dysfunction has been linked to cardiometabolic conditions which, in turn, increase the risk of depression.(52) For instance, elevated medium-chain ACs including octanoylcarnitine (C8) have been found(53) in gestational and Type-2 diabetes.

Beyond direct causal mechanisms, an association between AC metabolism and depression may arise from distal common environmental (e.g. exposure to stressful life events) or lifestyle (sedentariness, smoking, consumption of high-fat diet or alcohol) factors, which could also represent behavioral consequences of depression. Nevertheless, estimates derived in the present study via MR, suggesting a potential causal role of AC metabolism in the development of depression, are unlikely to be significantly biased by confounding factors. Furthermore, no evidence was found for a potential reversed causal role of depression in influencing AC levels. Other implicit properties of MR need to be taken into account when interpreting the present findings. Genetic variants index the average lifetime exposure to AC levels and, consequently, MR estimates derived from these instruments describe a potential average lifetime causal effect of ACs on depression and thus are unable to identify specific critical windows or acute events. Furthermore, since genetic instruments for depression were derived from large case-control GWAS, MR results provide information on potential causal mechanisms of disease onset rather than progression, which could involve different pathophysiological pathways. Finally, the causal relationship between ACs and depression identified in the present study requires confirmation across multiple causally-oriented methodologies (triangulation(54)), including further experimental and mechanistic studies in animals and humans.

A major strength of the present study is the application of 2SMR analyses leveraging novel GWAS summary statistics obtained from the largest international consortia. Nevertheless, despite the use of the largest available input data, the lack of statistically significant causal estimates may have resulted from insufficiently powered MR analyses, which may be determined by a combination of related factors, including the genetic architecture of the trait of interest, the sample size of the discovery GWAS, the number of independently associated SNPs and the strength of their associations. Biologically meaningful genetic instruments for ACs had adequate strength; this was confirmed in follow-up analyses in which stronger instruments derived from larger discovery GWAS summary statistics provided the same results as those obtained in the main analyses. Nevertheless, the number of independently associated genetic variants across ACs was low. In contrast, a higher number of genetic instruments for depression were available but are consistent with a polygenic architecture of weak effects scattered across the genome typical of all behavioral traits. Another limitation is that genetic instruments were derived from GWAS based on samples of European ancestry so results cannot be generalized to different populations. Thus, the present findings should be re-evaluated in the future when results of larger and more ancestry-diverse GWAS will become available. Finally, we could not properly deconvolute depression heterogeneity, which may aggregate different dimensions characterized by distinct pathophysiology. We previously showed (8,55) that inflammatory and metabolic alterations, potentially related to mitochondrial dysfunctions(56), maps more consistently to depressive “atypical” symptoms characterized by altered energy intake/expenditure balance (e.g hyperphagia, weight gain, hypersomnia, fatigue, leaden paralysis) and anhedonia. Future studies in large cohorts with deeper phenotypes should investigate whether the genetic signature of ACs is differentially associated with various depression clinical features and symptom profiles.

In conclusion, we found evidence of the potential causal role of AC metabolism on the risk of depression, in particular of high levels of medium-chain C8 and C10. Accumulation of medium-chain ACs is a signature of inborn errors of metabolism and age-related metabolic conditions. This suggests that altered cellular energy production and mitochondrial dysfunctions may have a role in depression pathogenesis. Acylcarnitine metabolism may represent a promising access point to depression pathophysiology suggesting novel therapeutic approaches.

## Data Availability

The present study is based on analyses applied to GWAS summary statistics that are available via the respective studies and consortia webportals.

https://omicscience.org/apps/crossplatform/

https://www.med.unc.edu/pgc/download-results/

https://dataverse.nl/dataset.xhtml?persistentId=doi:10.34894/JFWWS4

## Data Availability

https://omicscience.org/apps/crossplatform/

https://www.med.unc.edu/pgc/download-results/

https://dataverse.nl/dataset.xhtml?persistentId=doi:10.34894/JFWWS4

## Data Availability

https://omicscience.org/apps/crossplatform/

https://www.med.unc.edu/pgc/download-results/

https://dataverse.nl/dataset.xhtml?persistentId=doi:10.34894/JFWWS4

## Acknowledgments

YM and BWJHP acknowledge the following funding: this project has received funding from the European Union’s Horizon 2020 research and innovation programme under grant agreement No 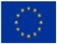 848146

In addition, the work was funded through NIMH grant number R01MH108348 and through a series of grants issued to Dr. Kaddurah-Daouk (PI) through NIA including U01AG061359, RF1AG057452, and RF1AG051550 that supported large number of scientists working on metabolomics and neuropsychiatric disorders and role for acylcarnitines.

M.A. and G.K. received funding (through their institutions) from the National Institutes of Health/National Institute on Aging through grants RF1AG058942, RF1AG059093, U01AG061359, U19AG063744, and R01AG069901.

The Fenland Study (10.22025/2017.10.101.00001) is funded by the Medical Research Council (MC_UU_12015/1). We further acknowledge support for genomics from the Medical Research Council (MC_PC_13046).

We are grateful to all Fenland volunteers and to the General Practitioners and practice staff for assistance with recruitment. We thank the Fenland Study Investigators, Fenland Study Co-ordination team and the Epidemiology Field, Data and Laboratory teams. We would like to thank the Psychiatric Genomics Consortium and 23andMe, including its research participants and employees, for making this work possible by sharing GWAS summary statistics.

## Disclousers

BWJHP has received research funding (unrelated to the work reported here) from Jansen Research and Boehringer Ingelheim.

A. John Rush has received consulting fees from Compass Inc., Curbstone Consultant LLC, Emmes Corp., Evecxia Therapeutics, Inc., Holmusk, Johnson and Johnson (Janssen), Liva-Nova, Neurocrine Biosciences Inc., Otsuka-US; speaking fees from Liva-Nova, Johnson and Johnson (Janssen); and royalties from Guilford Press and the University of Texas Southwestern Medical Center, Dallas, TX (for the Inventory of Depressive Symptoms and its derivatives). He is also named co-inventor on two patents: U.S. Patent No. 7,795,033: Methods to Predict the Outcome of Treatment with Antidepressant Medication, Inventors: McMahon FJ, Laje G, Manji H, Rush AJ, Paddock S, Wilson AS; and U.S. Patent No. 7,906,283: Methods to Identify Patients at Risk of Developing Adverse Events During Treatment with Antidepressant Medication, Inventors: McMahon FJ, Laje G, Manji H, Rush AJ, Paddock S.

M.A. and G.K. are co-inventors (through Duke University/Helmholtz Zentrum München) on patents on applications of metabolomics in diseases of the central nervous system; M.A. and G.K. hold equity in Chymia LLC and IP in PsyProtix and Atai that are exploring the potential for therapeutic applications targeting mitochondrial metabolism in depression.

S.M. is an inventor on patents in the Metabolomics field.

R.R.K. is CEO of Rush University System for Health, Chairman of National Medical Research Council Ministry of Health Singapore, Chairman of Amyriad, BV Board member Community Health Systems, Advisory board SageRx, Verily Patents on Brain Computer Interface licensed to Psyber and ATAI, an inventor of Metabolomics patents in the CNS field including patents licensed to Chymia LLC and PsyProtix with royalties and ownership. R.K.D. is funded by National Institute on Aging [U01AG061359, R01AG057452, RF1AG051550, and R01AG046171] and National Institute of Mental Health [R01MH108348]. This funding enabled consortia that she leads including the Mood Disorder Precision Medicine Consortium, the Alzheimer ’s disease Metabolomics Consortium, and the Alzheimer’s Gut Microbiome Project that contributed to acylcarnitine discoveries. She is an inventor on key patents in the field of Metabolomics and hold equity in Metabolon, a biotech company in North Carolina. In addition, she holds patents licensed to Chymia LLC and PsyProtix with royalties and ownership. The funders listed above had no role in the design and conduct of the study; collection, management, analysis, and interpretation of the data; preparation, review, or approval of the paper; and decision to submit the paper for publication.

B.W.D has received research support from Acadia, Compass, Aptinyx, NIMH, Sage, Otsuka, and Takeda, and has served as a consultant to Greenwich Biosciences, Myriad Neuroscience, Otsuka, Sage, and Sophren Therapeutics.

All the other authors declare no conflict of interest.

A preprint version of the manuscript has been deposited at medRxiv

(https://doi.org/10.1101/2021.10.18.21265157)

