## supplemental materials for "Genomics-based identification of a potential causal role for acylcarnitine metabolism in depression"

### **Supplemental methods**

#### ***MR-PRESSO***

The Mendelian randomization pleiotropy residual sum and outlier (MR-PRESSO) method(1) relies on a regression framework where the variants’ effects on the outcome are regressed on the same variants’ effects on exposure; the slope of the regression provides an estimate of the causal effect of the exposure on the outcome. The MR-PRESSO procedure includes several tests: 1) a global test evaluating overall horizontal pleiotropy among all instruments by comparing the observed residual sum of squares (RSS) with those expected under the null of no horizontal pleiotropy; 2) an outlier test evaluating the presence of specific outlier variants by using observed and expected distributions of the tested variants; 3) a distortion test evaluating the significance of the distortion in causal estimates before/after removal of outlier variants.

#### ***Follow-up analyses: GWAS meta-analysis***

In follow-up analyses evaluating the impact of the strength of genetic instruments on previous results, all 2SMR analyses were repeated for a subset of acylcarinites (ACs) with stronger genetic instruments derived from larger discovery GWAS summary statistic. For this we meta-analyzed results for the Fenland Study (2) with those from a previous GWAS meta-analysis by Draisma et al.(3) on 7,478 samples from 7 European cohorts with AC measures performed on the Biocrates AbsoluteIDQ p150 kit platform (Biocrates Life Sciences AG, Innsbruck, Austria). Genetic association results for 10 of the 15 metabolites examined in Fenland were available, including free carnitine (C0), short-chain (C2, C3), medium-chain (C8, C9, C10; C10:1) and long-chain (C14:1, C18:1, C18:2) ACs. These GWAS summary statics were pooled with those from the Fenland Study using a fixed-effects, inverse-weighted meta-analysis using METAL software(4). To address differences in the included sets of genetic variants due to differing imputation panels (HapMap phase 2 in Draisma et al., 1000 genomes phase 3 in Fenland Study), we first matched SNPs based on their identifiers and genomic locations. SNPs reported with differing alleles in Fenland and the Draisma et al. study were disregarded. In addition, AC levels were centered and scaled to zero mean and unit variance in the Fenland study but not in the Draisma et al. study, leading to inflated heterogeneity estimates in our meta-analysis of the two studies. To account for that, we rescaled genome-wide effect estimate distributions reported by Draisma et al. to the effect estimate distributions reported for Fenland, and centered standard error intervals to the thus derived, rescaled effect estimates. This led to substantially decreased, mostly insignificant heterogeneity across the vast majority of associations investigated in our meta-analysis. Exceptions were effect estimates at the extremes of the effect distributions (i.e., the most significant associations), which is expected from this statistical procedure. We therefore manually investigated genetic associations showing significant heterogeneity between the two studies. This identified the greater heterogeneity of the studies (n = 7 cohorts) underlying the initial Draisma et al. meta-analysis as the most likely cause, as it resulted overall in significantly lower reported effect estimates than those observed in the Fenland study, despite the similar sample sizes included in each of the studies.

In order to select genetic instruments for two-sample Mendelian randomization (2SMR) analyses, the pooled GWAS summary statistics were processed through similar steps as compared to those previously applied to Fenland Study results. AC summary statistics (~2M SNPs) were processed by removing strand ambiguous SNPs and with MAF<1%. Variants overlapping with those reported in depression GWAS were clumped (10,000 kb window, *r^2^*=0.01, EUR population of 1000Genomes used as linkage disequilibrium reference) to identify independent significantly (p<5.0e-8) associated SNPs. Genetic instruments selected for ACs and their strength is reported in eTable 9.

### **Supplemental Tables**

#### ***eTable 1.*** Selected genetic instruments for acylcarnitines

|  | | |  |  |  | **F-statistic** | |
| --- | --- | --- | --- | --- | --- | --- | --- |
| **Acylcarnitines** | | |  | **N SNPs** |  | min | max |
| *free carnitine* | Carnitine | C0 |  | 10 |  | 31.1 | 698.1 |
| *short-chain* | Acetylcarnitine | C2 |  | 5 |  | 31.2 | 297.1 |
|  | Propionylcarnitine | C3 |  | 9 |  | 29.8 | 326.9 |
|  | Butyrylcarnitine | C4 |  | 17 |  | 30.6 | 1312.6 |
|  | Valerylcarnitine | C5 |  | 5 |  | 32.2 | 160.8 |
| *medium-chain* | Hexanoylcarnitine | C6 |  | 6 |  | 31.3 | 348.2 |
|  | Octanoylcarnitine | C8 |  | 7 |  | 31.1 | 271.2 |
|  | Nonaylcarnitine | C9 |  | 2 |  | 48.1 | 275.3 |
|  | Decanoylcarnitine | C10 |  | 5 |  | 33.4 | 242.8 |
|  | Decenoylcarnitine | C10:1 |  | 5 |  | 30.0 | 128.7 |
|  | Dodecanoylcarnitine | C12 |  | 2 |  | 46.9 | 94.0 |
| *long-chain* | Tetradecenoylcarnitine | C14:1 |  | 2 |  | 35.6 | 55.1 |
|  | Hexadecanoylcarnitine | C16 |  | 2 |  | 59.3 | 70.3 |
|  | Octadecenoylcarnitine | C18:1 |  | 2 |  | 52.9 | 118.9 |
|  | Octadecandienylcarnitine | C18:2 |  | 2 |  | 32.8 | 88.1 |

F-statistic approximated as *est^2^/se^2^* according to Pierce et al.(5)

#### ***eTable 2.*** SNP-heritability estimates for acylcarnitines

|  |  | **h^2^_SNP_** | | |
| --- | --- | --- | --- | --- |
| **AC** |  | **est** | **se** | ***p-value*** |
| C0 |  | 0.12 | 0.07 | 7.59E-02 |
| C2 |  | 0.14 | 0.07 | 3.81E-02 |
| C3 |  | 0.15 | 0.06 | 1.84E-02 |
| C4 |  | 0.33 | 0.22 | 1.25E-01 |
| C5 |  | 0.21 | 0.09 | 1.40E-02 |
| C6 |  | 0.30 | 0.09 | 1.09E-03 |
| C8 |  | 0.25 | 0.08 | 2.08E-03 |
| C9 |  | 0.10 | 0.07 | 1.50E-01 |
| C10 |  | 0.21 | 0.07 | 1.80E-03 |
| C10:1 |  | 0.19 | 0.06 | 1.35E-03 |
| C12 |  | 0.18 | 0.05 | 2.60E-04 |
| C14:1 |  | 0.20 | 0.05 | 7.64E-05 |
| C16 |  | 0.11 | 0.06 | 7.48E-02 |
| C18:1 |  | 0.15 | 0.06 | 6.50E-02 |
| C18:2 |  | 0.14 | 0.05 | 9.43E-03 |

h^2^_SNP_ estimated using linkage-disequilibrium score regression (LDSC)(6)

#### ***eTable 3.*** Pairwise genetic correlations between acylcarnitines

| **ACs** | |  | ***rg*** | **se** | **p-value** |
| --- | --- | --- | --- | --- | --- |
| C2 | C2 |  | 0.14 | 0.07 | 3.81E-02 |
| C2 | C3 |  | 0.65 | 0.08 | 6.77E-15 |
| C2 | C5 |  | 0.62 | 0.19 | 1.41E-03 |
| C2 | C6 |  | 0.52 | 0.28 | 6.12E-02 |
| C2 | C8 |  | 0.43 | 0.19 | 2.28E-02 |
| C2 | C10 |  | 0.38 | 0.13 | 2.93E-03 |
| C2 | C10:1 |  | 0.39 | 0.1 | 6.00E-05 |
| C2 | C12 |  | 0.5 | 0.05 | 2.05E-27 |
| C2 | C14:1 |  | 0.68 | 0.05 | 6.42E-46 |
| C2 | C18:1 |  | 0.52 | 0.04 | 1.20E-40 |
| C2 | C18:2 |  | 0.26 | 0.04 | 1.27E-11 |
| C3 | C3 |  | 0.15 | 0.06 | 1.84E-02 |
| C3 | C5 |  | 0.78 | 0.09 | 1.95E-17 |
| C3 | C6 |  | 0.59 | 0.21 | 5.07E-03 |
| C3 | C8 |  | 0.42 | 0.12 | 3.85E-04 |
| C3 | C10 |  | 0.39 | 0.08 | 2.34E-06 |
| C3 | C10:1 |  | 0.31 | 0.11 | 4.76E-03 |
| C3 | C12 |  | 0.35 | 0.08 | 3.43E-05 |
| C3 | C14:1 |  | 0.39 | 0.14 | 7.11E-03 |
| C3 | C18:1 |  | 0.35 | 0.14 | 1.21E-02 |
| C3 | C18:2 |  | 0.29 | 0.06 | 1.31E-06 |
| C5 | C5 |  | 0.21 | 0.09 | 1.40E-02 |
| C5 | C6 |  | 0.59 | 0.23 | 9.55E-03 |
| C5 | C8 |  | 0.54 | 0.16 | 5.87E-04 |
| C5 | C10 |  | 0.54 | 0.07 | 1.22E-13 |
| C5 | C10:1 |  | 0.55 | 0.07 | 8.41E-16 |
| C5 | C12 |  | 0.55 | 0.11 | 1.02E-06 |
| C5 | C14:1 |  | 0.51 | 0.1 | 2.58E-07 |
| C5 | C18:1 |  | 0.53 | 0.09 | 2.21E-08 |
| C5 | C18:2 |  | 0.39 | 0.08 | 1.77E-06 |
| C6 | C6 |  | 0.3 | 0.09 | 1.09E-03 |
| C6 | C8 |  | 0.93 | 0.33 | 4.44E-03 |
| C6 | C10 |  | 0.9 | 0.26 | 6.80E-04 |
| C6 | C10:1 |  | 0.85 | 0.23 | 1.62E-04 |
| C6 | C12 |  | 0.69 | 0.31 | 2.51E-02 |
| C6 | C14:1 |  | 0.65 | 0.32 | 4.64E-02 |
| C6 | C18:1 |  | 0.58 | 0.28 | 3.88E-02 |
| C6 | C18:2 |  | 0.56 | 0.28 | 4.40E-02 |
| C8 | C8 |  | 0.25 | 0.08 | 2.08E-03 |
| C8 | C10 |  | 0.98^a^ | 0.01 | 2.32E-1133 |
| C8 | C10:1 |  | 0.95 | 0.21 | 8.25E-06 |
| C8 | C12 |  | 0.76 | 0.27 | 4.57E-03 |
| C8 | C14:1 |  | 0.64 | 0.27 | 1.63E-02 |
| C8 | C18:1 |  | 0.52 | 0.21 | 1.34E-02 |
| C8 | C18:2 |  | 0.56 | 0.22 | 9.95E-03 |
| C10 | C10 |  | 0.21 | 0.07 | 1.80E-03 |
| C10 | C10:1 |  | 0.98 | 0.17 | 3.61E-09 |
| C10 | C12 |  | 0.84 | 0.19 | 6.50E-06 |
| C10 | C14:1 |  | 0.66 | 0.18 | 3.16E-04 |
| C10 | C18:1 |  | 0.55 | 0.15 | 2.72E-04 |
| C10 | C18:2 |  | 0.61 | 0.21 | 4.30E-03 |
| C10:1 | C10:1 |  | 0.19 | 0.06 | 1.35E-03 |
| C10:1 | C12 |  | 0.87 | 0.14 | 7.02E-10 |
| C10:1 | C14:1 |  | 0.71 | 0.14 | 4.47E-07 |
| C10:1 | C18:1 |  | 0.6 | 0.13 | 4.28E-06 |
| C10:1 | C18:2 |  | 0.64 | 0.13 | 7.50E-07 |
| C12 | C12 |  | 0.18 | 0.05 | 2.60E-04 |
| C12 | C14:1 |  | 0.85 | 0.06 | 3.58E-52 |
| C12 | C18:1 |  | 0.57 | 0.08 | 4.31E-14 |
| C12 | C18:2 |  | 0.56 | 0.08 | 7.30E-12 |
| C14:1 | C14:1 |  | 0.2 | 0.05 | 7.64E-05 |
| C14:1 | C18:1 |  | 0.69 | 0.09 | 5.77E-16 |
| C14:1 | C18:2 |  | 0.53 | 0.08 | 1.68E-11 |
| C18:1 | C18:1 |  | 0.15 | 0.06 | 6.50E-02 |
| C18:1 | C18:2 |  | 0.69 | 0.11 | 6.31E-10 |
| C18:2 | C18:2 |  | 0.14 | 0.05 | 9.43E-03 |

*rg* estimates derived from high-definition likelihood (HDL)(7) and ^a^ bivariate LDSC(6) methods

#### ***eTable 4.*** Univariable Mendelian randomization analyses

|  |  |  |  | **Inverse Variance Weighted** | | | |  |
| --- | --- | --- | --- | --- | --- | --- | --- | --- |
| **Exposure** | **N SNPs** | **Outcome** |  | **OR** | **lCI** | **uCI** | ***p-value*** | ***q-value*** |
| C0 | 10 | DEP |  | 0.99 | 0.97 | 1.00 | 0.06 | 0.18 |
| C2 | 5 | DEP |  | 0.97 | 0.95 | 1.00 | 1.7E-02 | 0.06 |
| C3 | 9 | DEP |  | 0.97 | 0.96 | 0.99 | 6.1E-03 | 0.03 |
| C4 | 17 | DEP |  | 1.01 | 1.00 | 1.02 | 0.18 | 0.27 |
| C5 | 5 | DEP |  | 1.00 | 0.94 | 1.07 | 0.97 | 0.97 |
| C6 | 6 | DEP |  | 1.01 | 0.92 | 1.10 | 0.85 | 0.97 |
| C8 | 7 | DEP |  | 1.04 | 1.01 | 1.06 | 3.0E-03 | 0.02 |
| C9 | 2 | DEP |  | 1.02 | 0.99 | 1.05 | 0.16 | 0.27 |
| C10 | 5 | DEP |  | 1.04 | 1.02 | 1.06 | 6.7E-04 | 0.01 |
| C10:1 | 5 | DEP |  | 1.04 | 0.99 | 1.08 | 0.11 | 0.23 |
| C12 | 2 | DEP |  | 1.07 | 0.99 | 1.16 | 0.08 | 0.20 |
| C14:1 | 2 | DEP |  | 1.06 | 0.91 | 1.24 | 0.43 | 0.59 |
| C16 | 2 | DEP |  | 1.01 | 0.92 | 1.10 | 0.85 | 0.97 |
| C18:1 | 2 | DEP |  | 1.00 | 0.95 | 1.05 | 0.95 | 0.97 |
| C18:2 | 2 | DEP |  | 1.04 | 0.99 | 1.09 | 0.12 | 0.23 |

Odds ratios (ORs) and 95% confidence intervals (lCI, lower bound; uCI, upper bound) per SD increase in genetically-predicted levels of *_(log)_*ACs

*q*-value: false discovery rate (FDR) according to the Benjamini-Hochberg procedure.

#### ***eTable 5.*** Mendelian randomization sensitivity analyses: heterogeneity

|  |  |  |  | **Cochran's *Q*** | | |
| --- | --- | --- | --- | --- | --- | --- |
| **Exposure** | **N SNPs** | **Outcome** |  | ***Q*** | **df** | ***p-value*** |
| C2 | 5 | DEP |  | 1.34 | 4 | 0.85 |
| C3 | 9 | DEP |  | 4.46 | 8 | 0.81 |
| C8 | 7 | DEP |  | 12.66 | 6 | 0.05 |
| C10 | 5 | DEP |  | 5.28 | 4 | 0.26 |

#### ***eTable 6.*** Mendelian randomization sensitivity analyses: horizontal pleiotropy

|  |  |  |  |  |  |  |  | **MR-PRESSO** | | | | | | | | |
| --- | --- | --- | --- | --- | --- | --- | --- | --- | --- | --- | --- | --- | --- | --- | --- | --- |
|  |  |  |  | **MR-Egger intercept** | | |  | **Global Test** | |  | **Outlier corrected** | | |  | **Distortion Test** | |
| **Exposure** | **N SNPs** | **Outcome** |  | **intercept** | **se** | ***p-value*** |  | **RSS** | ***p-value*** |  | **est** | **se** | ***p-value*** |  | **Coeff** | ***p-value*** |
| C2 | 5 | DEP |  | 0.00003 | 0.00569 | 1.00 |  | 2.29 | 0.83 |  | n/a | n/a | n/a |  | n/a | n/a |
| C3 | 9 | DEP |  | -0.00477 | 0.00572 | 0.43 |  | 5.49 | 0.84 |  | n/a | n/a | n/a |  | n/a | n/a |
| C8 | 7 | DEP |  | 0.00332 | 0.00575 | 0.59 |  | 19.19 | 0.11 |  | n/a | n/a | n/a |  | n/a | n/a |
| C10 | 5 | DEP |  | 0.00005 | 0.00603 | 0.99 |  | 10.76 | 0.33 |  | n/a | n/a | n/a |  | n/a | n/a |

#### ***eTable 7.*** Reversed univariable Mendelian randomization analyses

|  |  |  |  | **Inverse Variance Weighted** | | | |
| --- | --- | --- | --- | --- | --- | --- | --- |
| **Exposure** | **N SNPs** | **Outcome** |  | **Estimate** | **lCI** | **uCI** | ***p-value*** |
| DEP | 96 | C0 |  | 0.003 | -0.086 | 0.092 | 0.95 |
| DEP | 96 | C2 |  | -0.026 | -0.115 | 0.063 | 0.57 |
| DEP | 96 | C3 |  | -0.021 | -0.106 | 0.064 | 0.62 |
| DEP | 96 | C4 |  | 0.081 | -0.140 | 0.301 | 0.47 |
| DEP | 96 | C5 |  | -0.020 | -0.110 | 0.070 | 0.66 |
| DEP | 96 | C6 |  | 0.049 | -0.039 | 0.137 | 0.27 |
| DEP | 96 | C8 |  | 0.006 | -0.082 | 0.095 | 0.89 |
| DEP | 96 | C9 |  | 0.032 | -0.063 | 0.127 | 0.51 |
| DEP | 96 | C10 |  | 0.018 | -0.071 | 0.106 | 0.70 |
| DEP | 96 | C10:1 |  | 0.002 | -0.086 | 0.089 | 0.97 |
| DEP | 96 | C12 |  | 0.008 | -0.081 | 0.097 | 0.86 |
| DEP | 96 | C14:1 |  | -0.055 | -0.144 | 0.035 | 0.23 |
| DEP | 96 | C16 |  | -0.002 | -0.099 | 0.095 | 0.96 |
| DEP | 96 | C18:1 |  | 0.009 | -0.080 | 0.098 | 0.84 |
| DEP | 96 | C18:2 |  | -0.005 | -0.103 | 0.092 | 0.91 |

Estimates and 95% confidence intervals (lCI, lower bound; uCI, upper bound) of change in SD of *_(log)_*AC levels per doubling (2-fold increase) in the prevalence of the exposure

#### ***eTable 8.*** Multivariable Mendelian randomization analyses

|  |  |  |  | **Multivariable MR** | | | |
| --- | --- | --- | --- | --- | --- | --- | --- |
| **Exposure** | **N SNPs** | **Outcome** |  | **OR** | **lCI** | **uCI** | ***p-value*** |
| C2 | 5 | DEP |  | 1.01 | 0.95 | 1.07 | 0.81 |
| C3 | 9 | DEP |  | 0.96 | 0.90 | 1.02 | 0.15 |
| C8 | 7 | DEP |  | 1.04 | 1.02 | 1.06 | 3.5E-05 |
| C2 | 5 | DEP |  | 0.99 | 0.95 | 1.04 | 0.80 |
| C3 | 9 | DEP |  | 0.97 | 0.93 | 1.02 | 0.19 |
| C10 | 5 | DEP |  | 1.04 | 1.02 | 1.06 | 5.6E-07 |
| C2 | 5 | DEP |  | 1.00 | 0.94 | 1.05 | 0.87 |
| C3 | 9 | DEP |  | 0.97 | 0.92 | 1.02 | 0.19 |
| C8 | 7 | DEP |  | 0.91 | 0.82 | 1.01 | 0.08 |
| C10 | 5 | DEP |  | 1.16 | 1.03 | 1.31 | 0.01 |

Odds ratios (ORs) and 95% confidence intervals (lCI, lower bound; uCI, upper bound) per SD increase in genetically-predicted levels of *_(log)_*ACs

#### ***eTable 9.*** Follow-up analyses: selected genetic instruments for acylcarnitines

|  |  | **F-statistic** | |
| --- | --- | --- | --- |
| **AC** | **N SNPs** | min | max |
| C0 | 9 | 33.1 | 1040.9 |
| C2 | 5 | 30.4 | 316.6 |
| C3 | 4 | 39.4 | 320.9 |
| C4 |  |  |  |
| C5 |  |  |  |
| C6 |  |  |  |
| C8 | 6 | 31.1 | 486.3 |
| C9 | 3 | 35.1 | 799.4 |
| C10 | 4 | 30.8 | 326.5 |
| C10:1 | 4 | 39.0 | 281.9 |
| C12 |  |  |  |
| C14:1 | 4 | 30.5 | 50.3 |
| C16 |  |  |  |
| C18:1 | 6 | 34.3 | 161.6 |
| C18:2 | 4 | 29.8 | 174.1 |

F-statistic approximated as *est^2^/se^2^* according to Pierce et al.(5)

Blank rows: AC not available in the GWAS meta-analyses by Draisma et al.(3)

#### ***eTable 10.*** Follow-up analyses: univariable Mendelian randomization analyses

|  |  |  | **Inverse Variance Weighted** | | | |
| --- | --- | --- | --- | --- | --- | --- |
| **Exposure** | **N SNPs** | **Outcome** | **OR** | **lCI** | **uCI** | ***p-value*** |
| C0 | 9 | DEP | 0.98 | 0.96 | 1.00 | 1.7E-02 |
| C2 | 5 | DEP | 0.96 | 0.94 | 0.99 | 1.4E-02 |
| C3 | 4 | DEP | 0.96 | 0.93 | 0.99 | 5.6E-03 |
| C4 |  |  |  |  |  |  |
| C5 |  |  |  |  |  |  |
| C6 |  |  |  |  |  |  |
| C8 | 6 | DEP | 1.05 | 1.02 | 1.08 | 2.7E-03 |
| C9 | 3 | DEP | 1.02 | 1.00 | 1.04 | 0.11 |
| C10 | 4 | DEP | 1.05 | 1.00 | 1.09 | 4.0E-02 |
| C10:1 | 4 | DEP | 1.06 | 0.99 | 1.12 | 0.09 |
| C12 |  |  |  |  |  |  |
| C14:1 | 4 | DEP | 1.06 | 0.94 | 1.19 | 0.34 |
| C16 |  |  |  |  |  |  |
| C18:1 | 6 | DEP | 1.02 | 0.98 | 1.07 | 0.33 |
| C18:2 | 4 | DEP | 1.02 | 0.96 | 1.09 | 0.48 |

Odds ratios (ORs) and 95% confidence intervals (lCI, lower bound; uCI, upper bound) per SD increase in genetically-predicted levels of *_(log)_*ACs

Blank rows: AC not available in the GWAS meta-analyses by Draisma et al.(3)

#### ***eTable 11.*** Follow-up analyses: multivariable Mendelian randomization analyses

|  |  |  |  | **Multivariable MR** | | | |
| --- | --- | --- | --- | --- | --- | --- | --- |
| **Exposure** | **N SNPs** | **Outcome** |  | **OR** | **lCI** | **uCI** | ***p-value*** |
| C0 | 9 | DEP |  | 0.98 | 0.92 | 1.04 | 0.45 |
| C2 | 4 | DEP |  | 1.04 | 0.90 | 1.20 | 0.61 |
| C3 | 5 | DEP |  | 0.95 | 0.85 | 1.07 | 0.44 |
| C8 | 6 | DEP |  | 1.05 | 1.02 | 1.07 | 1.4E-04 |
| C0 | 9 | DEP |  | 0.99 | 0.92 | 1.05 | 0.66 |
| C2 | 4 | DEP |  | 1.01 | 0.86 | 1.19 | 0.90 |
| C3 | 5 | DEP |  | 0.97 | 0.85 | 1.11 | 0.66 |
| C10 | 4 | DEP |  | 1.05 | 1.02 | 1.08 | 2.1E-03 |
| C0 | 9 | DEP |  | 0.98 | 0.93 | 1.04 | 0.51 |
| C2 | 4 | DEP |  | 1.03 | 0.90 | 1.17 | 0.69 |
| C3 | 5 | DEP |  | 0.96 | 0.87 | 1.07 | 0.46 |
| C8 | 6 | DEP |  | 0.90 | 0.85 | 0.95 | 0.51 |
| C10 | 3 | DEP |  | 1.18 | 0.99 | 1.41 | 0.06 |

Odds ratios (ORs) and 95% confidence intervals (lCI, lower bound; uCI, upper bound) per SD increase in genetically-predicted levels of *_(log)_*ACs

### **Supplemental figures**

#### ***eFigure 1.*** Mendelian randomization sensitivity analyses: single SNP analyses for Octanoylcarnitine (C8)

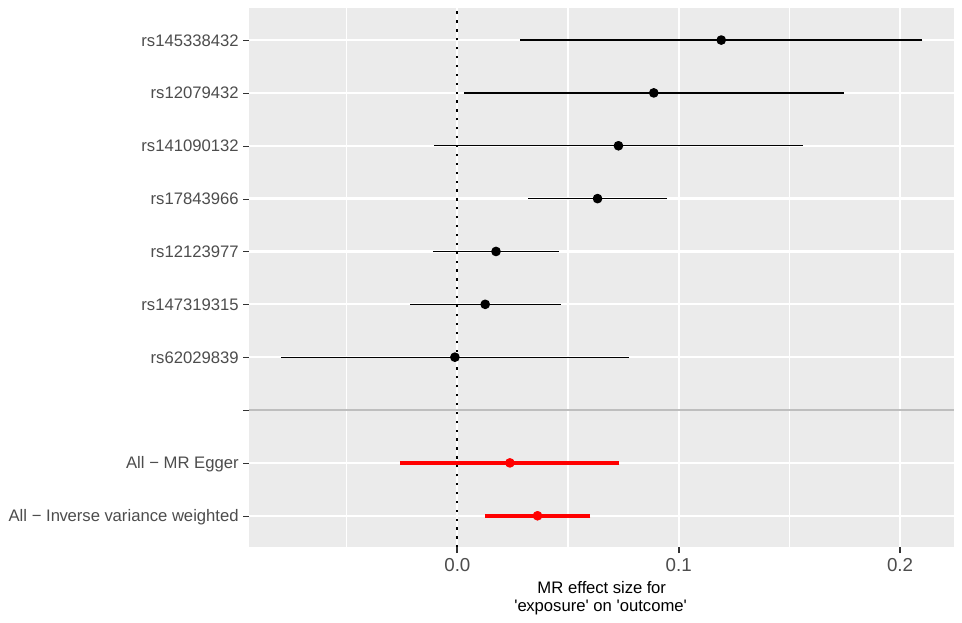

#### ***eFigure 2.*** Mendelian randomization sensitivity analyses: leave-one-out SNP analyses for Octanoylcarnitine (C8)

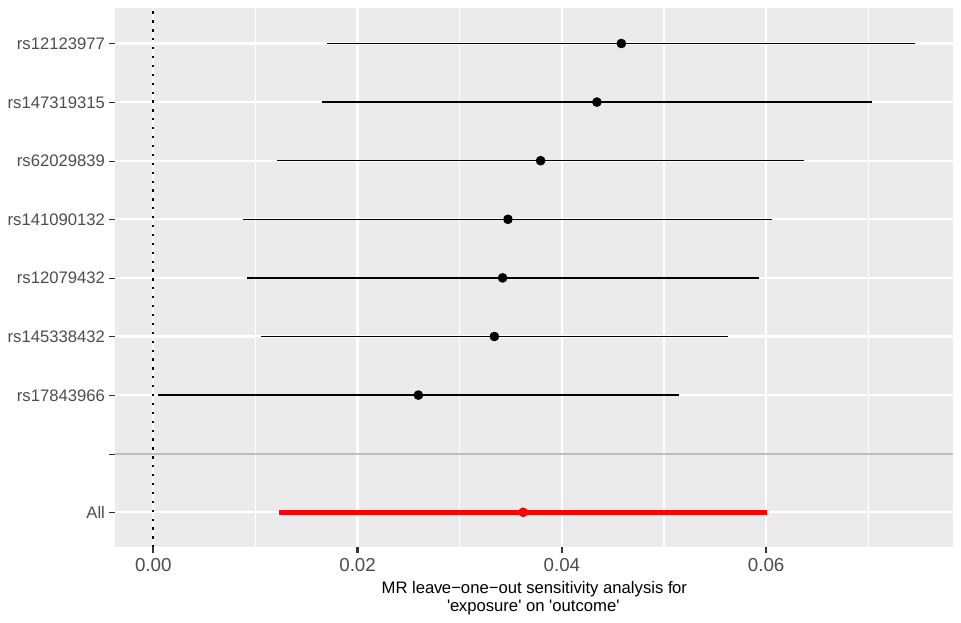

### **Data availability**

Summary statistics for ACs were retrieved from a GWAS in 9,363 samples from the Fenland study(2). GWAS summary statistics of genotype-metabolite associations are made available through an interactive web server (<https://omicscience.org/apps/crossplatform/>). Access to individual-level data can be requested at <https://epi-meta.mrc-epid.cam.ac.uk/>.

GWAS summary statistics for depression were obtained from the Psychiatric Genomics Consortium (PGC) overarching meta-analysis (8) of datasets with depression phenotypes, totaling 246,363 cases and 344,901 controls. The statistics publicly available here <https://www.med.unc.edu/pgc/download-results/> are based on a GWAS meta-analysis not including 23andMe data(9), access to which is restricted by a Data Transfer Agreement.

Full summary statistics for the metabolite GWAS meta-analysis by Draisma et al.(3) on 7,478 samples from seven European cohorts are available at <https://dataverse.nl/dataset.xhtml?persistentId=doi:10.34894/JFWWS4>
